# Theta, a multidimensional ratio biomarker applied to five amyloid beta peptides for investigations in familial Alzheimer’s disease

**DOI:** 10.1101/2025.08.06.25333131

**Authors:** Isaac Llorente-Saguer, Charles Arber, Neil P. Oxtoby

**Affiliations:** UCL Hawkes Institute and Department of Medical Physics and Biomedical Engineering, University College London, London, UK; Department of Neurodegenerative Disease, UCL Queen Square Institute of Neurology, University College London, London, UK; UCL Hawkes Institute and Department of Computer Science, University College London, London, UK

**Keywords:** Biomarker, amyloid, dimensionality reduction, classification, iPSC, PSEN1, Alzheimer’s disease, EOFAD

## Abstract

Biomarker discovery in complex diseases often requires classifying biological states using multiple, interconnected features, a task where traditional two-feature ratios are limited. We introduce theta, a mathematical model designed to extend the ratio concept for classification across any number of input features. Theta is a multivariate normative statistical model, which quantifies the deviation of a multidimensional feature vector relative to a reference. We apply theta to amyloid-beta (Aβ) peptide profiles to classify early-onset familial Alzheimer’s disease mutation carriers versus controls using multiple datasets of *in vitro* neuronal models. Theta consistently achieved high discrimination (AUC > 0.99, precision-recall AUC > 0.96) across multi-site datasets, considerably outperforming established Aβ biomarkers (precision-recall AUC 0.41 is the best score from the literature, in all datasets merged). This demonstrates theta’s power as a versatile mathematical tool.

## INTRODUCTION

Biomarker discovery in complex diseases, such as Alzheimer’s disease (AD), often relies on the identification of specific molecular changes that correlate with disease pathology or progression. While single analytes or simple ratios (i.e., A/B) have proven valuable, their utility can be limited by biological noise, confounding factors, or their inability to capture the subtle, interconnected shifts characteristic of heterogeneous pathological processes. We expand the concept of a ratio biomarker, extending it to any number of dimensions, instead of the classical two, in order to get a richer, more robust biological signal. This approach allows for leveraging the collective changes across an entire molecular profile, potentially offering superior diagnostic accuracy, prognostic power, and mechanistic insights compared to relying on simple two-component ratios.

Amyloid-beta (Aβ) peptides are an ideal showcase application for such a multidimensional analytical method. In AD, Aβ peptides are central to pathology, with their diverse forms and processing by-products reflecting the underlying disease state. Specifically, in both familial and sporadic AD, the selective deposition of longer Aβ peptides, particularly Aβ42, into brain amyloid plaques leads to a characteristic reduction in CSF Aβ42 levels, while Aβ40 levels remain relatively stable, thereby lowering the Aβ42/40 ratio^1^. In particular, mutations in *PSEN1*, linked to early-onset familial Alzheimer’s disease (EOFAD), profoundly impair Aβ precursor protein processing, resulting in a distinct molecular shift from shorter Aβ forms (Aβ37, Aβ38) to longer, aggregation-prone species (Aβ42, Aβ43)^2^. Notably, different mutations affect the Aβ profiles differently^3–5^. This complex landscape of Aβ isoforms and their dynamic changes underscores the need for comprehensive analytical approaches that capture the entire peptide profile rather than isolated ratios.

Early investigations have already hinted at the power of incorporating additional Aβ peptides beyond the canonical Aβ42/40 ratio. For instance, Liu and colleagues demonstrated the improved diagnostic utility of the Aβ37/42 ratio for AD, highlighting that even a two-peptide combination beyond Aβ42/40 could offer superior performance in cell models and CSF from sporadic AD patients^6^. Further, a previous work by Petit and colleagues introduced a composite ratio of (Aβ37+Aβ38+Aβ40)/(Aβ42+Aβ43) (termed short/long hereafter), which correlated with the age-at-onset of EOFAD mutations in cell models^7^. A recent study applied a data-driven model to investigate optimal linear combinations of peptide ratios^8^, and also found a composition similar to that of Petit et al. These pioneering studies collectively underscore the principle that a more comprehensive integration of multiple Aβ peptide profiles may offer superior diagnostic and prognostic power, providing a rationale for developing formal mathematical models to achieve this.

The clinical translation of AD biomarkers increasingly favours minimally invasive, cost-effective, and scalable blood-based assays. Historically, blood-based Aβ peptide ratios faced challenges in achieving diagnostic accuracy comparable to CSF measures due to lower concentrations and confounding factors^9^. However, recent advancements in highly sensitive analytical techniques, such as single-molecule array (Simoa)^10^, Nucleic acid linked immuno-sandwich assay (NULISA^™^)^11^, immunoprecipitation-mass spectrometry (IP-MS)^12^, have dramatically improved the performance of plasma Aβ42/40 and Aβ42/40/p-tau ratios. These modern assays now demonstrate considerable diagnostic utility for detecting brain amyloidosis, often comparable to CSF and PET measures in identifying individuals with underlying AD pathology^13,14^. While these breakthroughs have validated the utility of established blood-based ratios, there is a critical and unmet need to apply more sophisticated, multidimensional analytical frameworks to Aβ data to capture subtle nuances, improve precision for specific aetiologies like EOFAD, and potentially offer earlier detection or prognostic insights.

Given the established molecular parallels between iPSC-derived neuronal Aβ profiles and blood plasma ratios^15^, we sought to leverage our previously established iPSC-derived neuronal model of familial AD^8^ as a robust platform for biomarker development. In this study, we design and propose a mathematical model aimed at deriving multidimensional ratios, and apply it to investigate a comprehensive panel of Aβ peptides.

## RESULTS

In this study, theta is our proposed multidimensional ratio biomarker, which integrates peptide measures into a single composite number. We performed classification experiments on multiple in vitro datasets — and on the merged dataset — using two complementary metrics: the area under the receiver operating characteristic curve (AUC) for overall discrimination and the area under the precision-recall curve (PRAUC) to robustly assess performance on the minority class (controls). As benchmarks, we compared theta against established literature-derived Aβ ratio biomarkers (Aβ42/40, Aβ37/42, and (Aβ37+38+40)/(Aβ42+43)) to evaluate its relative effectiveness.

Figure 1. visually represents the separation of theta scores between control and EOFAD samples, per dataset. The theoretical best theta value for controls is 0 (an angle of 0 radians); the four closest mutations to the controls, in theta, are: E318G (theta = 0.053), considered benign^16,17^; A164V (theta = 0.074), with unclassified^18^ pathogenicity; E69D (theta = 0.082), with unclassified pathogenicity^17,19^; D40del (delGAC) (theta = 0.088), with uncertain^20^ pathogenicity. The apparent visual separation will be quantitatively assessed in the next paragraphs.

**Figure 1:**
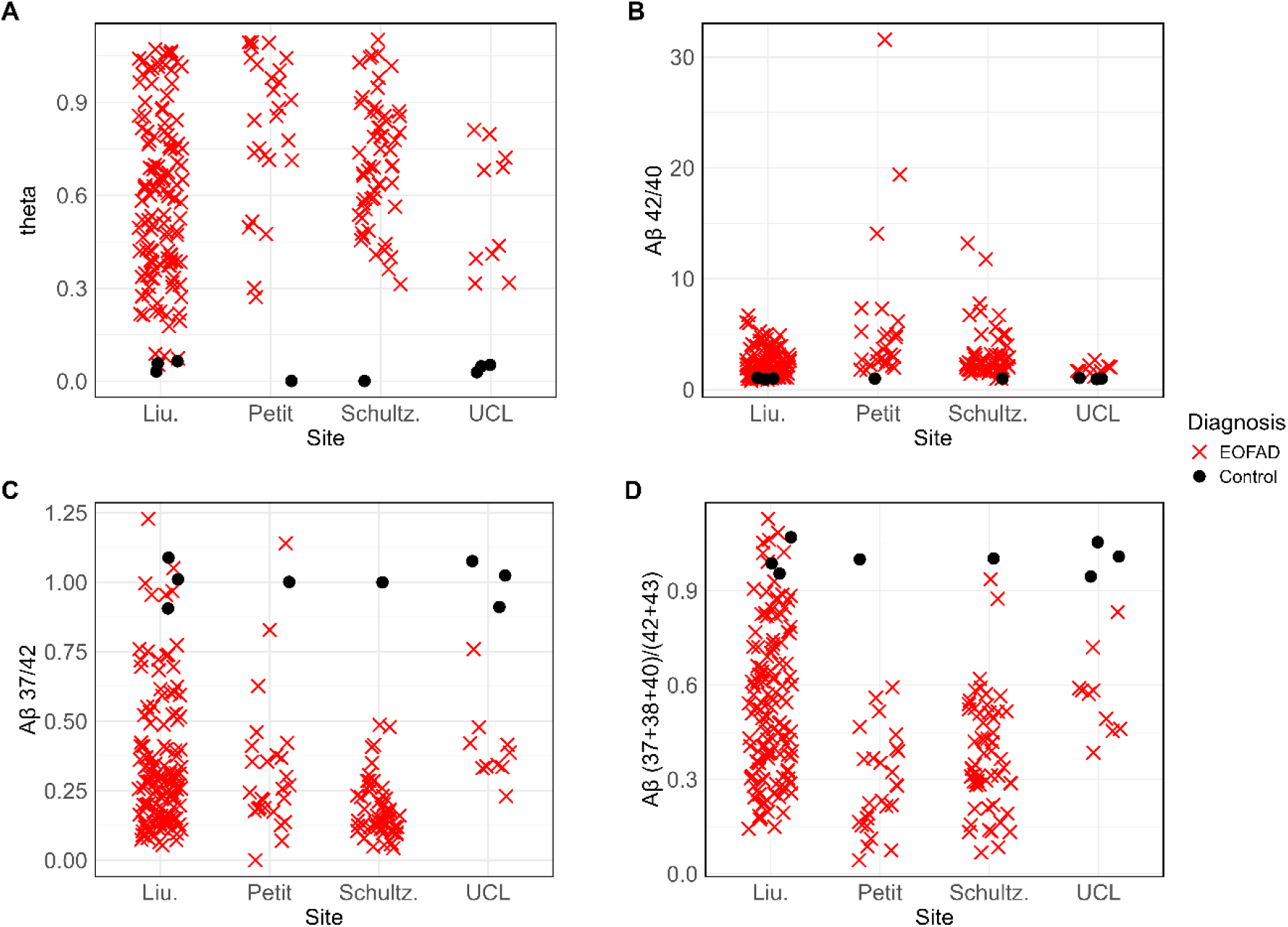
Biomarker values per site. Black circles represent controls; red crosses represent PSEN1 mutations.

**Table 1** summarises the overall classification performance (controls versus EOFAD PSEN1 mutations) of each biomarker across the merged datasets, with 95 % confidence intervals obtained via 5000 stratified bootstrap samples. All biomarkers achieved very high AUCs under the tightly controlled experimental conditions, but performance diverged on the more stringent precision–recall metric focused on the minority (control) class. Specifically, our theta-biomarker achieved an AUC of 0.999 (95 % CI 0.992– 1.000) and a PR AUC of 0.964 (95 % CI 0.661–1.000), substantially outperforming the literature benchmarks. The best literature biomarker was Aβ37/42, with an AUC of 0.984 and PR AUC of 0.410 in all merged datasets. When restricting to double-knockout cell data only, theta maintained similarly superior performance (AUC 0.999, 95 % CI 0.993–1.000; PR AUC 0.969, 95 % CI 0.827–1.000), whereas the other biomarkers saw marked drops in precision–recall despite still high AUCs.

**Table 1:** Classification evaluation in merged datasets, with 95% confidence intervals by 5000 stratified bootstrapping samples. Data are sourced from four studies: Llorente et al., Liu et al., Petit et al., and Schultz et al. We evaluated the biomarkers merging all data, and all double-knockout cell data (all data except Llorente et al.). In bold, the highest performance metrics. AUC = Area under the receiver operating characteristic curve (true positive rate versus false positive rate). PR AUC = Area under the precision-recall curve, focused on the minority class in the data: the controls.

| Biomarker | AUC | PR AUC |
| --- | --- | --- |
| **All double-knockout cell data** |  |  |
| 42/40 | 0.960 (0.929, 0.983) | 0.291 (0.187, 0.564) |
| 37/42 | 0.984 (0.965, 0.997) | 0.501 (0.312, 0.935) |
| (37+38+40)/(42+43) | 0.976 (0.953, 0.993) | 0.395 (0.254, 0.743) |
| theta | 0.999 (0.993, 1.000) | 0.969 (0.827, 1.000) |
| **All data merged** |  |  |
| 42/40 | 0.956 (0.921, 0.982) | 0.201 (0.122, 0.438) |
| 37/42 | 0.984 (0.963, 0.997) | 0.410 (0.227, 0.919) |
| (37+38+40)/(42+43) | 0.975 (0.950, 0.992) | 0.299 (0.179, 0.661) |
| theta | 0.999 (0.992, 1.000) | 0.964 (0.661, 1.000) |

Pairwise comparisons (**Table 2**) confirmed that theta is significantly better than every other biomarker on both PR AUC and ROC AUC in the merged dataset (all p < 0.01), with the strongest differences against the weakest performer (Aβ42/40; p < 0.0001). Among the non-theta markers, only the 37/42 ratio consistently outperformed 42/40 (0.005 < p < 0.04).

**Table 2:**
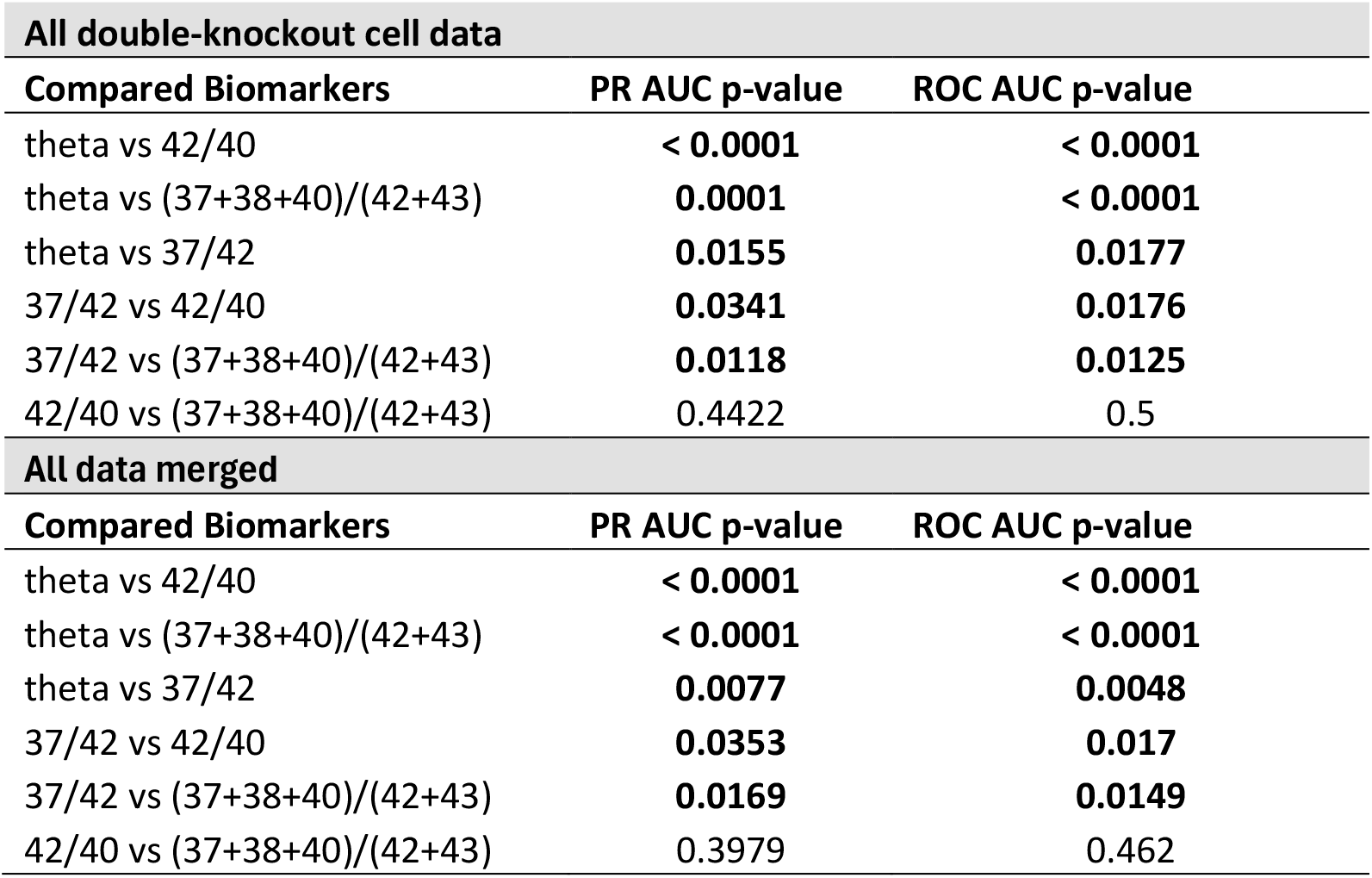
Pairwise comparisons of biomarker performance metrics. P-values (uncorrected) were derived from 10,000 stratified bootstrap samples for each biomarker pair to test whether the row biomarker outperforms the column biomarker, in merged datasets; values (uncorrected) below 0.05 indicate significant differences (shown in bold).

| All double-knockout cell data |  |  |
| --- | --- | --- |
| Compared Biomarkers | PR AUC p-value | ROC AUC p-value |
| theta vs 42/40 | < 0.0001 | < 0.0001 |
| theta vs (37+38+40)/(42+43) | 0.0001 | < 0.0001 |
| theta vs 37/42 | 0.0155 | 0.0177 |
| 37/42 vs 42/40 | 0.0341 | 0.0176 |
| 37/42 vs (37+38+40)/(42+43) | 0.0118 | 0.0125 |
| 42/40 vs (37+38+40)/(42+43) | 0.4422 | 0.5 |
| All data merged |  |  |
| Compared Biomarkers | PR AUC p-value | ROC AUC p-value |
| theta vs 42/40 | < 0.0001 | < 0.0001 |
| theta vs (37+38+40)/(42+43) | < 0.0001 | < 0.0001 |
| theta vs 37/42 | 0.0077 | 0.0048 |
| 37/42 vs 42/40 | 0.0353 | 0.017 |
| 37/42 vs (37+38+40)/(42+43) | 0.0169 | 0.0149 |
| 42/40 vs (37+38+40)/(42+43) | 0.3979 | 0.462 |

Site-specific analyses (**Table 3**) revealed heterogeneity in classification difficulty. In the Llorente et al. cohort—all biomarkers including theta reached perfect separation under those conditions—but in the Liu et al. dataset, theta still achieved near-perfect discrimination (AUC 0.995, 95 % CI 0.980–1.000; PR AUC 0.851, 95 % CI 0.389–1.000), whereas the benchmarks fell to PR AUCs between 0.161 and 0.389. Petit and Schultz cohorts exhibited patterns similar to Llorente (perfect scores) and Liu (variable PR AUC), indicating that the non-control group is not from a single uniform class, but rather an aggregation of multiple different PSEN1 mutations. Hence the need to aggregate multicohort data for more generalisable interpretation of biomarker performance.

**Table 3:**
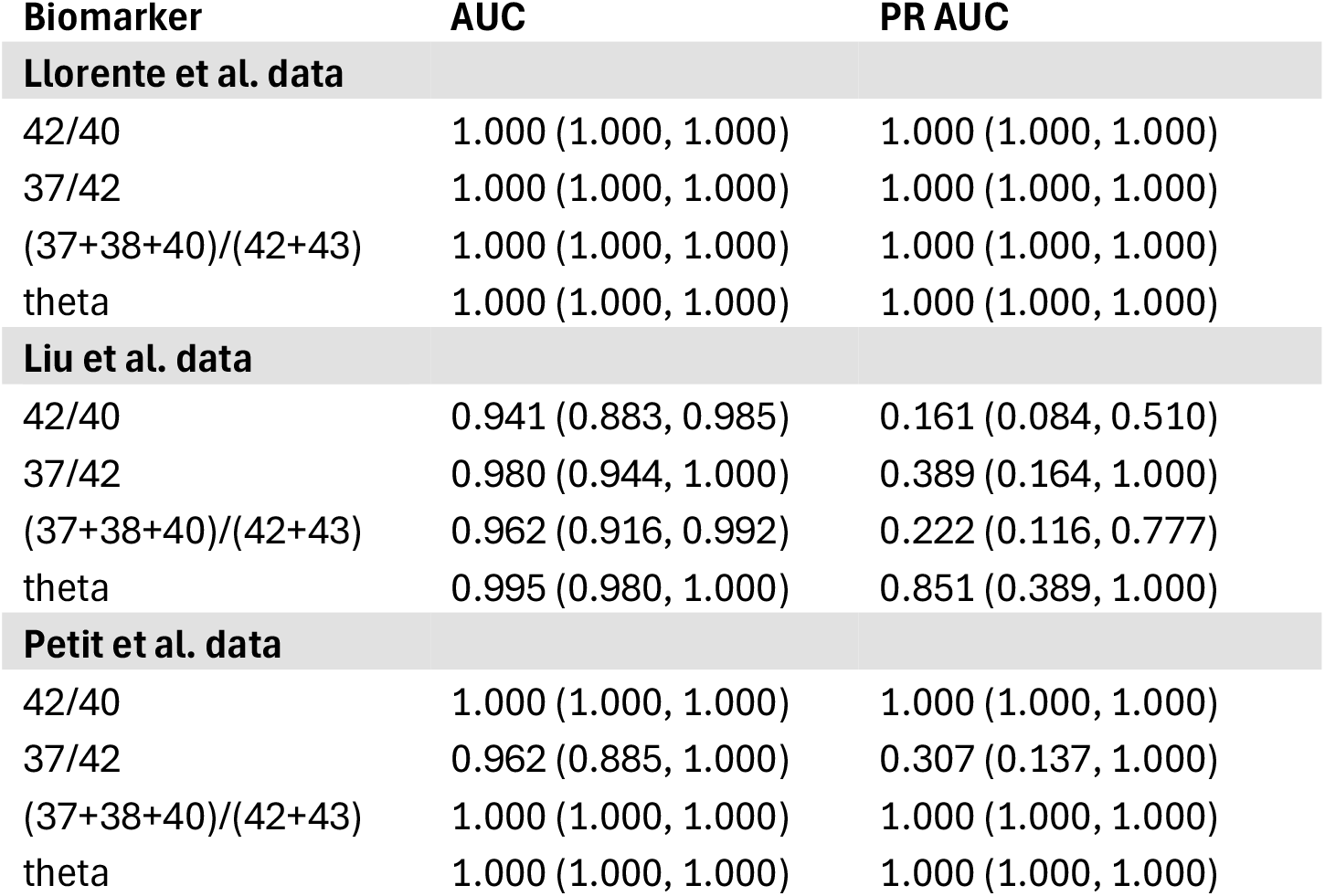
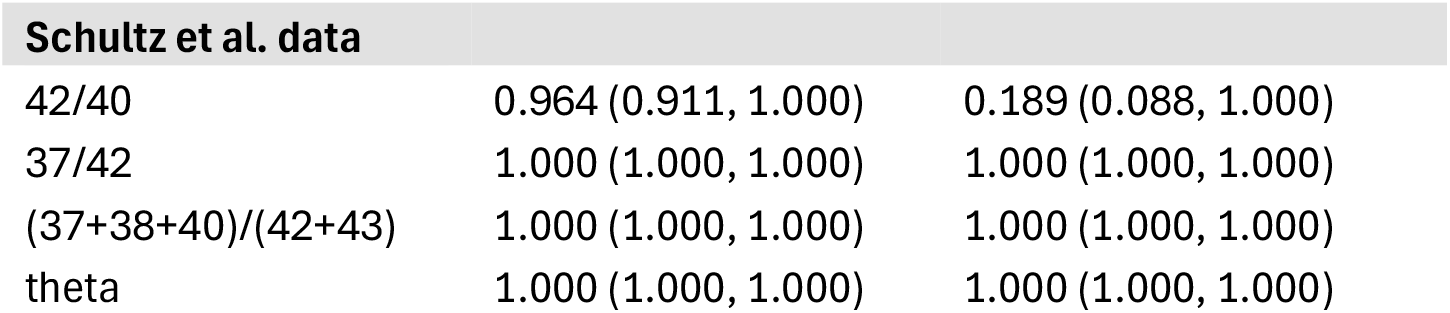
Classification evaluation in specific datasets, with 95% confidence intervals by 5000 stratified bootstrapping samples. AUC = Area under the receiver operating characteristic curve (true positive rate versus false positive rate). PR AUC = Area under the precision-recall curve, focused on the minority class in the data: the controls. Data comes from four sources: Llorente et al., Liu et al., Petit et al. and Schultz et al.

| Biomarker | AUC | PR AUC |
| --- | --- | --- |
| Llorente et al. data |  |  |
| 42/40 | 1.000 (1.000, 1.000) | 1.000 (1.000, 1.000) |
| 37/42 | 1.000 (1.000, 1.000) | 1.000 (1.000, 1.000) |
| (37+38+40)/(42+43) | 1.000 (1.000, 1.000) | 1.000 (1.000, 1.000) |
| theta | 1.000 (1.000, 1.000) | 1.000 (1.000, 1.000) |
| Liu et al. data |  |  |
| 42/40 | 0.941 (0.883, 0.985) | 0.161 (0.084, 0.510) |
| 37/42 | 0.980 (0.944, 1.000) | 0.389 (0.164, 1.000) |
| (37+38+40)/(42+43) | 0.962 (0.916, 0.992) | 0.222 (0.116, 0.777) |
| theta | 0.995 (0.980, 1.000) | 0.851 (0.389, 1.000) |
| Petit et al. data |  |  |
| 42/40 | 1.000 (1.000, 1.000) | 1.000 (1.000, 1.000) |
| 37/42 | 0.962 (0.885, 1.000) | 0.307 (0.137, 1.000) |
| (37+38+40)/(42+43) | 1.000 (1.000, 1.000) | 1.000 (1.000, 1.000) |
| theta | 1.000 (1.000, 1.000) | 1.000 (1.000, 1.000) |
| **Schultz et al. data** |  |  |
| 42/40 | 0.964 (0.911, 1.000) | 0.189 (0.088, 1.000) |
| 37/42 | 1.000 (1.000, 1.000) | 1.000 (1.000, 1.000) |
| (37+38+40)/(42+43) | 1.000 (1.000, 1.000) | 1.000 (1.000, 1.000) |
| theta | 1.000 (1.000, 1.000) | 1.000 (1.000, 1.000) |

## DISCUSSION

Our study introduced a novel multidimensional biomarker ratio (theta) and applied it to Aβ peptide data for classifying early-onset familial Alzheimer’s disease (EOFAD) mutations from control samples in various iPSC-derived neuronal models. We hypothesised that integrating signal across multiple Aβ isoforms would detect the subtle, coordinated shifts of AD pathology more reliably than single-ratio measures. The consistent high discrimination achieved by theta across multiple independent and merged datasets validated this hypothesis, marking a step forward in biomarker development for complex neurological disorders.

The superior classification performance of theta versus benchmark biomarker ratios in these *in vitro* iPSC-derived neuronal models is noteworthy. These models, directly derived from patients, faithfully recapitulate key aspects of human Aβ pathology driven by specific genetic mutations, offering a highly controlled environment to isolate precise molecular signatures. In this context, the underlying biological shifts in Aβ processing in EOFAD, as captured by our multidimensional ratio, appear highly distinct and consistently measurable. This sharply contrasts with benchmark biomarkers (Aβ42/40, Aβ37/42, and the “short/long” ratio), which, although frequently demonstrating high AUCs (often inflated by considerable class imbalance), displayed considerably lower performance in precision-recall (PRAUC). This discrepancy highlights the inherent limitations of simple ratios in fully capturing the complex biological signal when multiple Aβ species are perturbed. Theta maintained high precision and recall, even amid class imbalance, which underscores its robustness and potential to generate reliable diagnostic signals, suggesting its utility for detecting even subtle pathological deviations or in cases where genetic causes are less clear or unknown.

We emphasise several essential aspects to comprehend the mechanistic basis of theta’s exceptional performance: first and foremost, the nature of the data is such that single peptide measures can be affected by experimental variables (e.g., concentration of the solution, cultivation time and conditions), so it is imperative to use a ratio to cancel out these factors. This translates into a direction in the multidimensional space of the analysed features, where there is no contribution, just noise (e.g., multiplying all peptide measures by a scalar will not change their relative abundance). Theta removes this dimension and keeps only how much the vectorised input features’ direction deviates from the default direction of the controls. Second, as reported in previous studies, different mutations affect peptides differently^8^. Third, data from controls is understood to possess a specific profile, rather than being heterogeneous. Thus, theta moves beyond simple one-dimensional ratios, leveraging the comprehensive Aβ molecular profile to effectively integrate the diverse effects of genetic mutations on Aβ production and processing. This provides a compelling example of how multivariate approaches can extract more informative signals from biological data, surpassing the capabilities of metrics that use fewer variables. In essence, theta captures the deviation of any input variable with respect to a normative profile, which becomes highly useful when abnormalities can occur in multiple variables.

Despite the strong performance in *in vitro* models, a critical next step involves validating theta’s performance in human biospecimens, specifically CSF and plasma from EOFAD patients, sporadic AD patients, and appropriate control cohorts. Blood‐based AD markers are advancing fast^21^, and theta’s multidimensional readout of plasma peptides (amyloid and tau) could be a timely approach to boost precision. Translational challenges include the inherent variability in human biological samples, the influence of peripheral Aβ metabolism, and the need for highly sensitive and standardised analytical platforms capable of accurately quantifying the full range of Aβ peptides (Aβ37, Aβ38, Aβ40, Aβ42, Aβ43) in blood.

Our mathematical model for theta is designed to integrate information from all available Aβ features to maximise its classification power. Although this approach can be advantageous — when each feature contributes useful, potentially distinct discriminatory power — it also means the model considers every input. This can become a limitation if a particular input feature is excessively noisy or less informative. Future work can easily extend the theta concept to incorporate methods for identifying the most impactful subset of features or to explicitly quantify and include feature importance weightings. Such refinements could potentially simplify future assays while maintaining high performance and provide deeper insights into the key driving molecular changes.

While this study focuses on EOFAD, the principles underlying theta’s multidimensional design could be adapted to identify more sensitive biomarkers for sporadic AD, other neurodegenerative diseases, or even for assessing therapeutic efficacy. The mathematical model developed here offers a flexible framework that can be applied to other panels of related biomarkers or features, extending its utility beyond Aβ peptides to other protein isoforms, metabolites, or even multi-omic data. In practice, this means that a researcher could substitute or augment Aβ with neurofilament light chain (NFL) to quantify axonal injury^22,23^, α-synuclein species to probe synucleinopathies^24,25^, or the spectrum of 3R/4R tau isoforms to resolve tauopathy subtypes^26^, without altering the mathematical formulation of the model. The same logic extends to markers such as GFAP^27,28^ and sTREM2^29^ that capture inflammatory states/neuronal injury.

In conclusion, our new multidimensional ratio-based biomarker theta, applied here on AB peptide data demonstrated unparalleled diagnostic accuracy for EOFAD in iPSC-derived neuronal models. This shows the value of integrating complex molecular information through mathematical modelling to generate robust and precise biomarkers. While further validation in human cohorts is essential, theta potentially represents a significant conceptual and practical advancement in biomarker discovery, offering a promising avenue for improving the diagnosis, prognosis, and potentially the monitoring of Alzheimer’s disease and other complex human pathologies.

## METHODS

### Data

The data used for the analysis in this work consist of measurements of Aβ peptides from iPSC and cell cultures from multiple cohorts, as described in Table 4. To account for assay and cohort differences, each peptide measurement was normalised by dividing it by the mean value of the corresponding control group for that peptide and cohort^8^.

**Table 4:** Data from different sources, with the number of controls and PSEN1 mutation-carriers. Specific mutations are given in each respective source.

| Dataset | Controls | PSEN1 |
| --- | --- | --- |
| Llorente et al. 2024, BioRxiv<sup>8</sup> | 3 | 10 |
| Liu et al. 2023, *Alzheimers Dement.*<sup>6</sup> | 3 | 131 |
| Petit et al. 2022, *Mol. Psychiatry*<sup>7</sup> | 1 | 26 |
| Schultz et al. 2024, *Lancet Neurol.*<sup>30</sup> | 1 | 56 |
| **Total** | **8** | **223** |

### Theta

Each data sample consists of five peptide measures, which can be represented as a point in a five-dimensional space. Simple ratio biomarkers, such as Aβ42/40 and Aβ37/42, work in the projection of the multidimensional space onto a plane defined by the two selected peptides (see **Figure 2**). The peptide ratio is the tangent of the angle formed between the vector from the origin to the data point and the axis of the denominator peptide dimension (**Figure 3**). Since peptide measures cannot be negative, all possible data points lie within the positive orthant and, therefore, this angle is a monotonic function of the ratio, as is its complementary angle.

**Figure 2:**
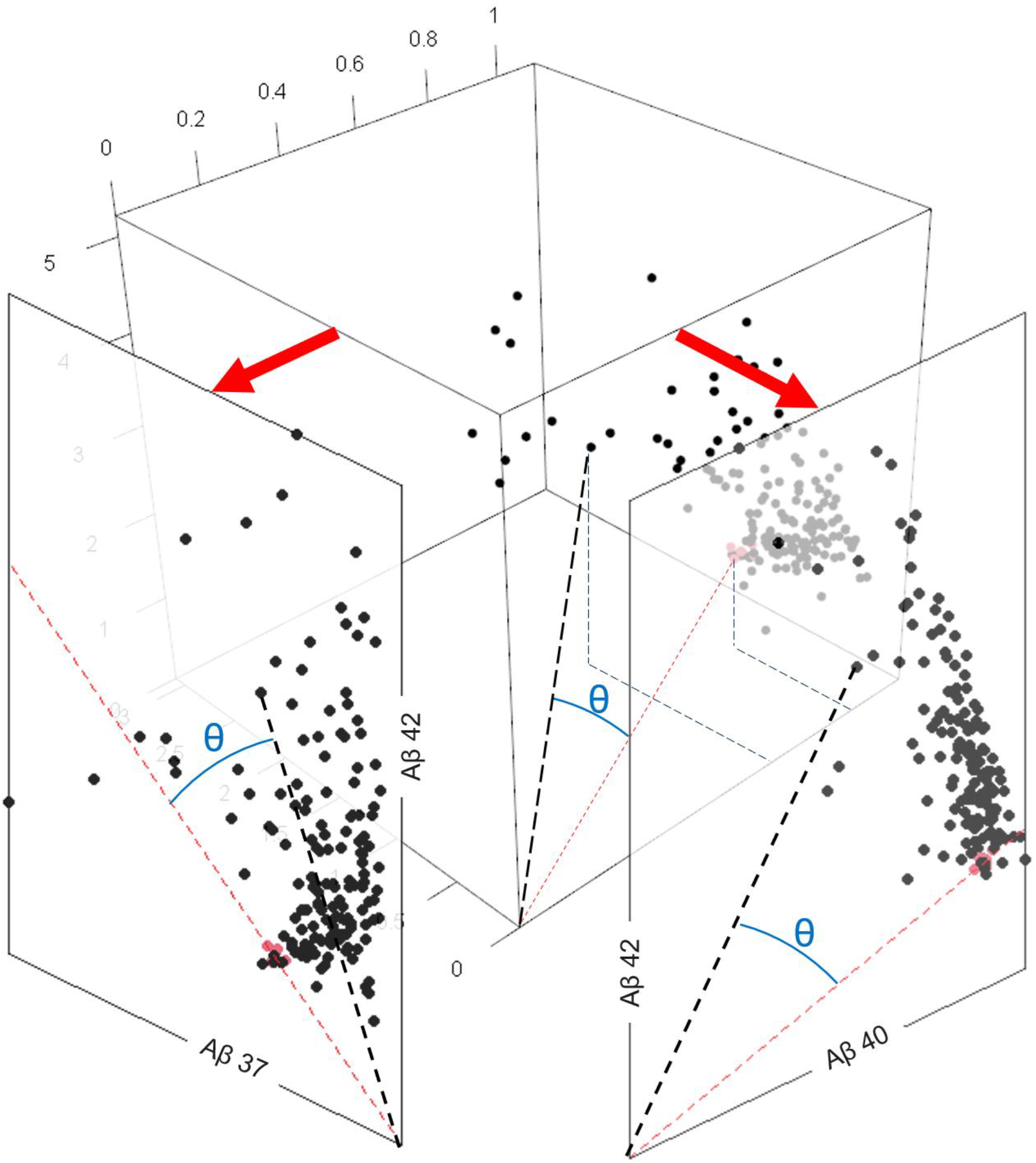
Distribution of the three merged cell datasets in three dimensions, corresponding to Aβ 37, 40 and 42. Red dots represent controls. This is a graphical representation of ratio usage. When using simple ratio peptides like Aβ 42/40 and Aβ 37/42 for classification, users will essentially compare the direction of the controls (red dashed line) with respect to that of a datapoint (black). We define theta as the angle between these two directions, which can be in any number of dimensions (e.g., three in this figure, or two).

**Figure 3:**
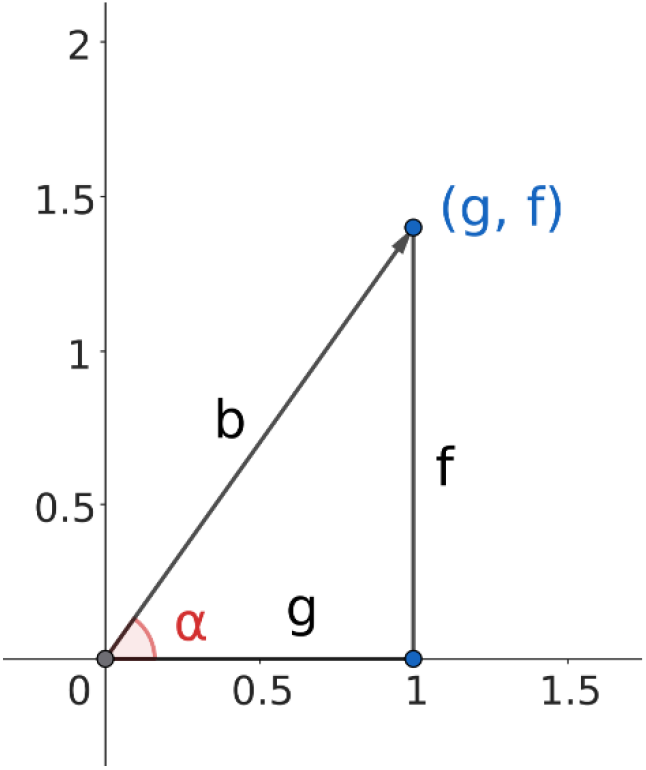
Relationship between the components of a ratio (f/g) and the angle of the vector (b) they form, with respect to the numerator. The ratio f/g is equal to the tangent of the angle alpha. Figure created in GeoGebra (geogebra.org).

When using simple two-variable ratio peptides like Aβ 42/40 and Aβ 37/42 for classification, users will essentially compare the direction of the controls (red dashed line in **Figure 2**) with respect to that of a datapoint (black dashed line in **Figure 2**). We define theta as the angle between these two directions, which can be in any number of dimensions. After all peptide measures are divided by the average of the controls in each dataset as a harmonisation step, all controls are centred around the direction defined by the vector-of-ones 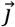 We can then use this direction as a reference, and calculate the angle θ with respect to any other origin-datapoint vector 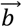 (**Figure 2**). Using the cosine similarity, calculating the angle theta (θ) is straightforward, as shown in the following equation:

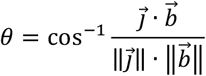

where the numerator is the dot product of vectors 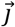 *and* 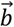, and the denominator is the multiplication of the magnitudes (norms) of vectors 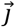 and 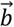, respectively. Note that due to the data harmonisation, this method can be used in any dataset without any fitting of parameters required.

### Benchmarks

We compare the performance of theta to Aβ42/40^31,32^, Aβ37/42^6^ and “short/long” = (Aβ37+38+40)/(Aβ42+43)^7^.

### Evaluation metrics

Given the heavy class imbalance in the data (with relatively few controls), we utilise the precision-recall curve (PR AUC) for the smaller group, which plots controls’ precision TN/(TN+FN) against controls’ recall TN/(TN+FP), where T/F denotes True/False and P/N signifies Positive/Negative. To align with the literature, we also provide the widely used AUC (area under the receiver operating characteristic curve). We provide 95% confidence intervals using 5000 bootstrapped samples stratified by class (controls and mutation-carriers). The significance of metric differences will be assessed using stratified bootstrapping (10000 samples).

All statistical analyses are performed using R version 4.5.0^33^.

## Data Availability

All data produced in the present study are available upon reasonable request to the authors.

## RESOURCE AVAILABILITY

### Lead contact

Further information and requests for resources and reagents should be directed to and will be fulfilled by the lead contact, Neil Oxtoby.

### Materials availability

The data from Llorente et al.^8^ is pending publication in a peer-reviewed journal, upon which the data will be available to other researchers. The rest of the data utilised in this study comes from open-access literature and is available for other researchers to use.

### Data and code availability

Code to calculate theta (in R and Python) is available at the following repository: github.com/isaac-6/theta

## ACKNOWLEDGMENTS

I.Ll.S. and N.P.O. acknowledge funding from a UKRI Future Leaders Fellowship (MR/S03546X/1, MR/X024288/1). C.A. was supported by Race Against Dementia and the National Institute for Health and Care Research University College London Hospitals Biomedical Research Centre. We also acknowledge the work of the respective datasets used in this study.

## AUTHOR CONTRIBUTIONS

Conceptualization, I.Ll.S.; methodology, I.Ll.S.; writing—original draft, I.Ll.S.; writing—review & editing, N.P.O. and C.A.; funding acquisition, N.P.O.

## DECLARATION OF GENERATIVE AI AND AI-ASSISTED TECHNOLOGIES

During the preparation of this work, ILLS used Grammarly to improve the grammar. After using this tool or service, the authors reviewed and edited the content as needed and take full responsibility for the content of the publication.

